# Molecular genetic study on *HAND2* gene promoter in ventricular septal defect

**DOI:** 10.1101/2022.06.28.22277011

**Authors:** Meikun Li, Yahui Cai, Shuchao Pang, Bo Yan

**Affiliations:** Cheeloo College of Medicine, Shandong University, Jinan, Shandong, China; Institute of Precision Medicine, Jining Medical University, Jining, Shandong, China; College of Basic Medicine, Jining Medical University, Jining, Shandong, China; Shandong Provincial Sino-US Cooperation Research Center for Translational Medicine, Affiliated Hospital of Jining Medical University, Jining Medical University, Jining, Shandong, China

## Abstract

Ventricular septal defect (VSD), as the most common type of congenital heart disease (CHD), is mainly caused by cardiac dysplasia. *Heart and neural crest derivatives expressed 2* (*HAND2)*, as a member of the *basic helix-loop-Helix (bHLH)* gene family, participates in the development of the right heart. And the loss of *HAND2* expression in humans is closely connected with ventricular septal defects. However, no studies have hitherto examined the association between VSD and *HAND2* gene promoter. We used a case-control study to analyze the genetic variations of the *HAND2* gene promoter region in VSD patients and controls. Using a variety of statistical analysis methods to analyze the association of single nucleotide polymorphisms (SNPs) with VSD. The dual luciferase reporter assay was used to conduct functional analysis of genetic variations. Electrophoretic mobility shift assay (EMSA) was used to examine DNA-protein interactions. Through sequencing, we identified nine genetic variants in patients with VSD. The SNP rs2276940 G>T and rs2276941 G>A were revealed to have associations with an increased risk of VSD. The dual luciferase reporter assay showed that the SNP rs2276940 G>T and rs138531627 C>G decreased the transcriptional activity of the *HADN2* gene promoter. Transcription factors (TFs) predicting suggested that all three above SNPs may change the binding of the TFs. The result of EMSA showed that rs138531627 C>G may create a new binding site for TFs while rs2276940 G>T enhanced the binding affinity for TFs. All these results indicated that genetic variants of the *HAND2* gene promoter increase VSD risk by reducing *HAND2* expression, and the molecular mechanism may be the change of the binding affinity of TFs.

## Introduction

To date, congenital heart defect (CHD) is still the most common congenital anomaly, as well as a leading cause of infant death from birth defects [1-3]. Despite advances in diagnostics and surgery, there is still a high neonatal mortality rate on account of congenital heart defect [4]. Multiple later complications such as heart failure, endocarditis and arrhythmias are the leading causes of death[3]. Ventricular septal defect (VSD) is the most common type of congenital heart disease [5]. The same as other types of congenital heart defect, the major pathogenesis of ventricular septal defect is cardiac dysplasia[6].

The development of the heart, the earliest functional organ formed in the mammalian embryo, requires the precisely control of transcriptional regulation networks and cell signaling pathways[7, 8]. During embryonic heart development, cardiac progenitor cells in the mesoderm differentiate into the first heart field and the second heart field, in which the first heart field forms the left ventricle and the left and right atrium, and the second heart field forms the right ventricle, outflow tract and part of the atrium[9]. Multiple cardiac transcription factors (TFs) such as *GATA binding protein* (*GATA)*[10], *heart and neural crest derivatives expressed* (*HAND)*[11], NK2 homeobox 5 (*Nkx2–5)*[12], and the *T-box family members*[13] play critical roles in this process. As a matter of course, mutations in these TFs will contribute to cardiac dysplasia, as well as lead to VSD[14].

The *HAND* gene, as a member of the *basic helix-loop-Helix (bHLH)* gene family, is highly conserved[15]. It has two isoforms that are asymmetrically expressed in developing ventricular chambers, namely *HAND1* and *HAND2*. Before the formation of the heart tube, *HAND1* and *HAND2* are jointly expressed in the lateral mesoderm and neural crest. With the development of the heart, *Hand2* is concentrated in the second heart field, and finally specifically expressed in the right ventricle and outflow tract, while as *HAND1* is mainly expressed in the left ventricle[16, 17]. The stimulative role of *HAND2* in the development of cardiomyocytes came from the second heart field was suggested in vivo, as loss of *HAND2* in mice leads to hypoplasia of the right ventricle and outflow tract[17]. And the increase of the expression levels of *HAND2* was found in patients with right ventricular hypertrophy[18]. Further experiment confirms that loss of *HAND2* in the second heart field will enhance apoptosis of cardiac progenitor cells before they take part in the process of heart development, contributing to serious hypoplasia of the right ventricle[19]. Simultaneously, an increasing body of evidence suggests that abnormal expression of *HAND2* in humans is closely connected with congenital heart defect that include ventricular septal defects[20, 21], despite recent study suggesting that *HAND2* is a specific factor for outflow tract cells rather than right ventricular cells[22].

Based on the above studies, we consider that the loss of *HAND2* expression is one of the leading causes contributing to ventricular septal defect. However, the regulatory network of *HAND2* and the molecular mechanism that leads to the decrease of *HAND2* expression during the development of ventricular septal defect are still unknown. Therefore, in this study, we attempted to identify the genetic variants on the *HAND2* promoter that leads to the decrease of *HAND2* expression and then induces ventricular septal defect formation, and further explore its molecular mechanism.

## Materials and methods

### Study participants

VSD patients (n=349; 204 males and 145 females) were recruited from Division of Cardiac Surgery, Affiliated Hospital of Jining Medical University, Jining, Shandong, China from December 2018 to February 2021. VSD patients were diagnosed according to clinical manifestation, physical examination, electrocardiogram and three-dimensional echocardiography. Ethnic-matched healthy controls (n=345, 168 males, 177 females) were recruited from Physical Examination in the same hospital during the same time period. VSD patients who had structural malformations involving another organ system or familial CHD history were excluded. This study strictly followed the principles of the Helsinki Declaration (1964) and was approved by the Human Ethic Committee of Affiliated Hospital of Jining Medical University. Written informed consent was obtained from all participants.

### Sequence analysis

Peripheral leukocytes were isolated using the Human Leukocyte Isolation system (Haoyang Biological Products Technology Co., Ltd., Tianjin, China). Genomic DNAs were extracted using the QIAamp DNA Mini kit (Qiagen, Inc., Valencia, CA, United States). The *HAND2* promoter region from -1300 to +46 bp (1346 bp, chr4: 173530184-173531529, NC_000004.12) was amplified using PCR. Using BLAST program on NCBI to design the PCR primers [Forward: 5’-(Kpn I)-GTTGACACCGTTTTCCACACC-3’; Reverse: 5’-(Hind III)-TGAGGGGAGCAAGCGGATTT-3’]. The PCR products were sequenced by Sangon Biotech Co., Ltd. (Shanghai, China), and compared with the wild-type *HAND2* gene promoter using the DNAMAN program.

### Functional analysis by dual-luciferase reporter assay

DNA fragments of wild type and variant *HADN2* gene promoters were generated by PCR using the above primers, and then subcloned into Kpn I and Hind III sites of vector (pGL3-basic) to generate expression vectors. Designated expression vectors and expression vector pRL-TK were transiently transfected into primary neonatal rat cardiomyocytes (NRCMs) and human embryonic kidney cells (HEK-293) (CRL-1573; ATCC, Manassas, VA, USA). Using dual-luciferase reporter assay system on a Glomax 20/20 luminometer (Promega, Madison, WI, USA) to measure the luciferase activity of the transfected cells after 48 h. Expression vector pRL-TK was taken as an internal control. *HAND2* gene promoter activity was showed as the ratio of luciferase activity over Renilla luciferase activity. Empty vector pGL3-basic was used as a negative control. The transcription activity of the wild-type *HAND2* gene promoter was set as 100%. All the experiments were repeated three times independently, in triplicate.

### Isolation of primary neonatal rat cardiomyocytes (NRCMs)

Two SPF-grade SD rats, purchased from Jinan Pengyue Experimental Animal Breeding Co., LTD., the license number is SCXK (Lu) 2019-0003. Neonatal rat pups (1–3 days old, gender unknown) from the mother were disinfected with 75% alcohol before removing their hearts. The extracted heart was put into PBS containing 100 U/mL penicillin streptomycin to remove non cardiac tissue and residual blood, and then cut the heart into pieces. The chopped tissue was digested with 0.25% trypsin for 5 minutes, and then the supernatant was transferred to the complete medium (15% fetal bovine serum and 100 U/mL penicillin-streptomycin). Repeat the above steps until the tissue is completely digested. The supernatant was filtered into a new EP tube using a cell strainer and then centrifuged (1000 rpm, 15 min). The centrifuged cells were collected into a petri dish, added with complete medium, and then cultured at 37°C in a 5% CO_2_ incubator for 1.5 hours. Collected the supernatant and planted it into the 6-well plate. Transfection was performed after 36 to 48 hours[23, 24].

### Electrophoretic mobility shift assay

NE-PER® Nuclear Protein/Cytoplasmic Protein Extraction Kit (Thermo Fisher Scientific, Inc.) was used to prepare nuclear extracts of the HEK-293 and NRCMs cells. The protein concentration of the nuclear extract was measured using Bio-Rad Protein Assay Reagent and stored at –80°C. Biotinylated double-stranded oligonucleotides (30 bp) containing genetic variants were used as probes. The DNA-protein binding reaction to explore the interaction between the DNA fragment of HAND2 gene promoter and the nucleoprotein was conducted using LightShift® chemiluminescence electrophoretic mobility shift assay (EMSA) kit (Thermo Fisher Scientific, Inc.).

### Statistical analysis

Quantitative data were expressed as means ± standard deviation (SD) or means ± standard error of mean (SEM) and analyzed using a Student’s t-test. The qualitative data were analyzed and compared by chi-square test. Each genetic variants frequency in the VSD and control subjects was tested for deviation from Hardy–Weinberg equilibrium (HWE) by the Fisher’s test. The above analyses were conducted using SPSS 22.0 statistical software packages. The web-based software SNP-Stats (https://www.snpstats.net/start.htm) was used to analyse the association between five genetic models (codominant, dominant, over-dominant, recessive, and log-additive) and VSD, and the results were showed as odds ratio (OR) and 95% confidence interval (CI). P < 0.05 was considered as statistically significant.

## Results

### Nine genetic variants were identified in patients with VSD

A total of twelve genetic variants were identified in the promoter of *HAND2*, through sequencing in 349 VSD patients and 345 healthy control subjects (Fig 1). The VSD patients included 204 males (58.5%) and 145 females (41.5%), and the mean age was 5.39±2.85 years. The controls included 168 males (48.7%) and 177 females (51.3%), and the mean age was 5.40±2.76 years. There was no significant difference in age between the two groups (P = 0.49). Compared with controls, VSD patients included more males (P = 0.01). Nine genetic variants, including eight single nucleotide polymorphisms (SNPs), were identified in patients with VSD (Fig 2A) and another three were present only in controls (Fig 2B). The chi-square test showed that the distribution of g.173530848 G>T (rs2276940) and g.173530503 G>A (rs2276941) in VSD patients and healthy control subjects are statistically significant (Table 1). The Online website SNPStats was used to analyse the correlations between multiple genetic models of the above two SNPs and AMI. The genotype distribution of the SNP rs2276940 G>T conforms to the codominant genetic model, dominant genetic model, recessive genetic model and overdominant genetic model. While the genotype distribution of the SNP rs2276941 G>A conforms to the dominant genetic model and overdominant genetic model (Table 2). Linkage disequilibrium (LD) analysis showed that the two SNPs (rs2276940 and rs2276941) are no in obvious linkage disequilibrium (D ‘=0.9064, r^2^=0.1988). The haplotype analysis showed the most significant risk effect for the HAND2 haplotype rs2276940T/rs2276941A (P=0.0035) (Table 3).

**Fig 1.**
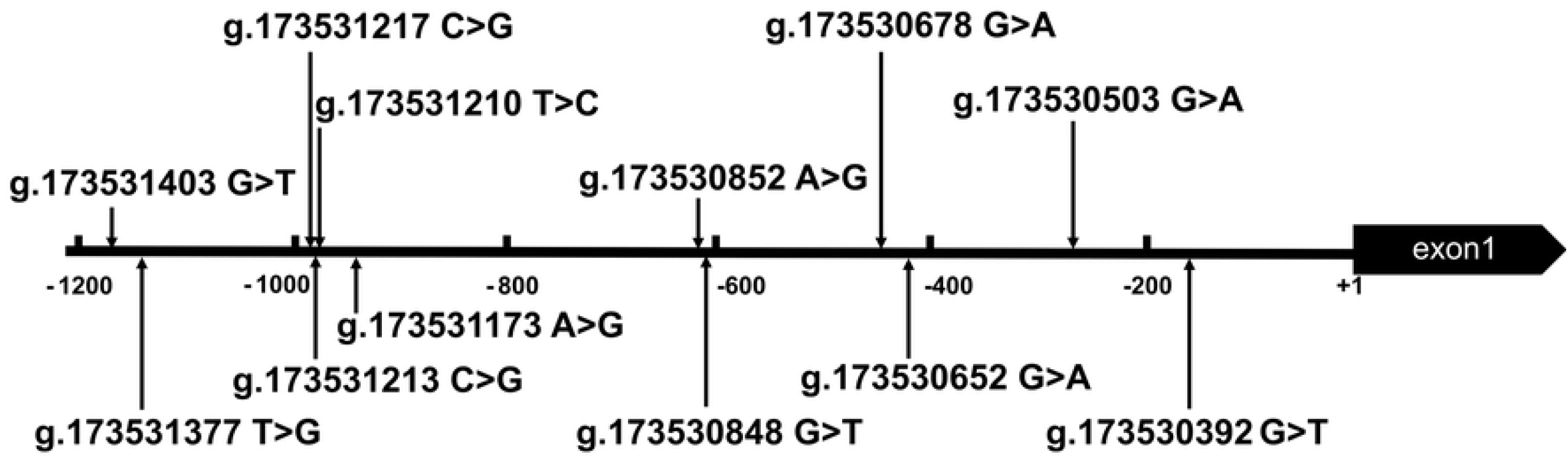
Locations of genetic variants on *HAND2* gene promoter. The numbers represent the genomic DNA sequences of the *HAND2* gene (NC_000004.12) upstream to the transcription start site (+1).

**Fig 2.**
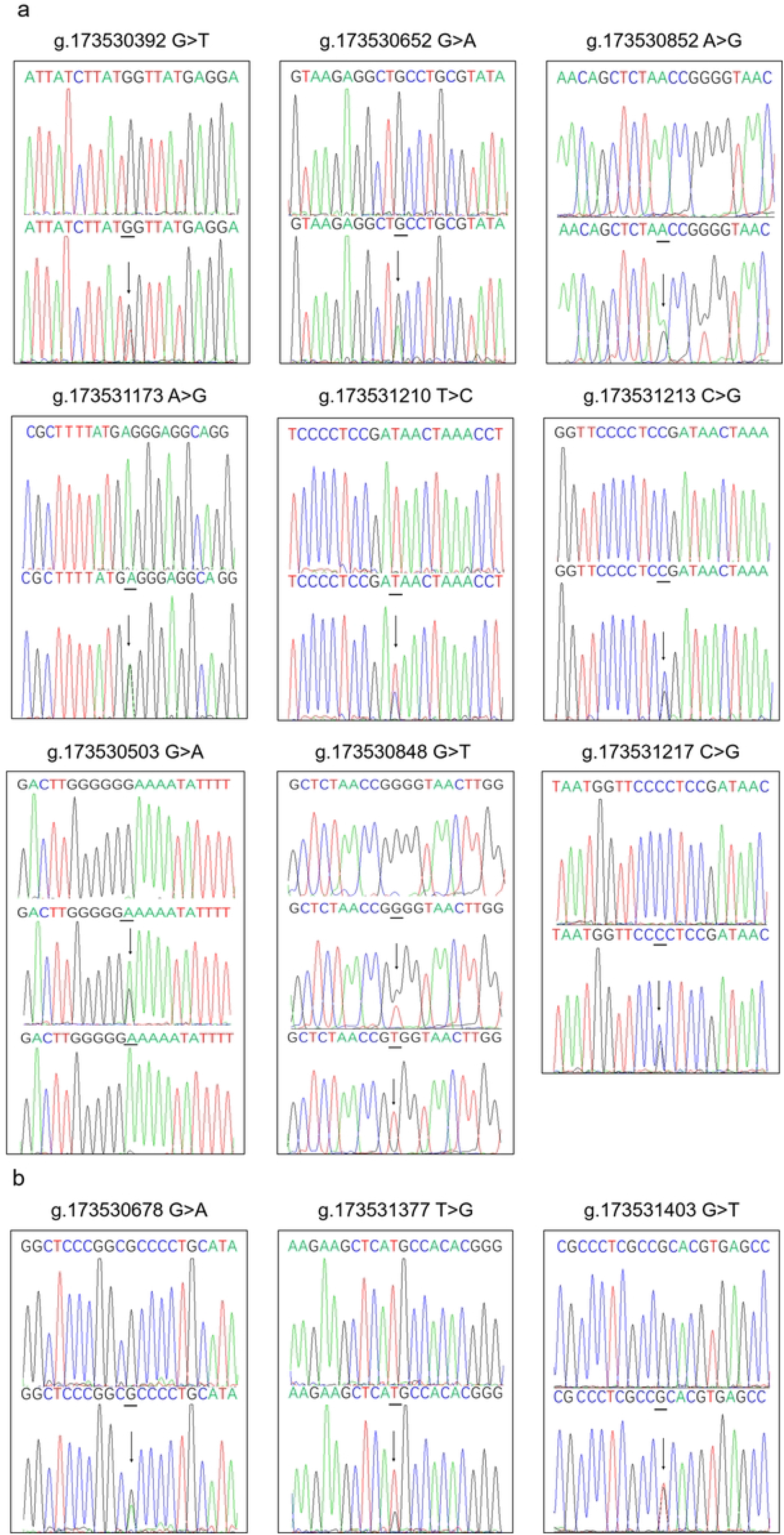
Sequencing chromatograms of the genetic variants on *HAND2* gene promoter. (a). Sequencing chromatograms of the genetic variants in VSD patients. (b). Sequencing chromatograms of the genetic variants in healthy controls. All sequence orientations of the genetic variants are forward. The top layers are wild-type sequences, the bottom panels are variants, and the variant sites are marked with arrows.

**Table 1.** Genetic variants of *HAND2* gene promoter in VSD patients and controls.

| genetic variants | Genotype | Location* | Control(n=345) | VSD(n=349) | P value |
| --- | --- | --- | --- | --- | --- |
| g.173531403 G>T (rs532631502) | GT | -1174bp | 1 | 0 | - |
| g.173531377 T>G | TG | -1148bp | 1 | 0 | - |
| g.173531217 C>G (rs187801237) | CG | -988bp | 3 | 8 | 0.134 |
| g.173531213 C>G (rs138531627) | CG | -984bp | 0 | 7 | - |
| g.173531210 T>C (rs1313594565) | TC | -981bp | 1 | 1 | 0.993 |
| g.173531173 A>G | AG | -944bp | 0 | 1 | - |
| g.173530852 A>G (rs552312676) | AG | -623bp | 0 | 2 | - |
| g.173530848 G>T (rs2276940) | GG | -619bp | 316 | 298 | 0.017 |
|  | GT |  | 29 | 48 |  |
|  | TT |  | 0 | 3 |  |
| g.173530678 G>A(rs1731703893) | GA | -449bp | 1 | 0 | - |
| g.173530652 G>A(rs1360489005) | GA | -423bp | 2 | 1 | 0.556 |
| g.173530503 G>A(rs2276941) | GG | -274bp | 227 | 196 | 0.031 |
|  | GA |  | 112 | 147 |  |
|  | AA |  | 6 | 6 |  |
| g.173530392 G>T(rs371582671) | GT | -163bp | 2 | 3 | 0.663 |

\* : variants are located upstream to the transcription start site of *HAND2* gene at g.173530229 (NC\_000004.12).

**Table 2.** Genetic models’ analysis of the association between the SNPs in the *HAND2* promoter region and VSD (adjusted by SEX + AGE).

| SNP | Model | Genotype | Control | VSD | OR (95%CI) | P value | HWP* |
| --- | --- | --- | --- | --- | --- | --- | --- |
| rs2276940 | Codominant | G/G | 317 (91.90%) | 299 (85.70%) | 1.00 | 0.012 | 1.000 |
|  |  | G/T | 28 (8.10%) | 47 (13.50%) | 1.72 (1.05-2.83) |  |  |
|  |  | T/T | 0 (0.00%) | 3 (0.90%) | - |  |  |
|  | Dominant | G/G | 317 (91.90%) | 299 (85.70%) | 1.00 | 0.014 |  |
|  |  | G/T-T/T | 28 (8.10%) | 50 (14.30%) | 1.83 (1.12-3.00) |  |  |
|  | Recessive | G/G-G/T | 345 (100.00%) | 346 (99.10%) | 1.00 | 0.045 |  |
|  |  | T/T | 0 (0.00%) | 3 (0.90%) | - |  |  |
|  | Overdominant | G/G-T/T | 317 (91.90%) | 302 (86.50%) | 1.00 | 0.032 |  |
|  |  | G/T | 28 (8.10%) | 47 (13.50%) | 1.71 (1.04-2.80) |  |  |
|  | Log-additive | - | - | - | 1.87 (1.17-3.01) | 0.007 |  |
| rs2276941 | Codominant | G/G | 227 (65.80%) | 196 (56.20%) | 1.00 | 0.071 | 0.068 |
|  |  | G/A | 112 (32.50%) | 147 (42.10%) | 1.45 (1.06-1.98) |  |  |
|  |  | A/A | 6 (1.70%) | 6 (1.70%) | 1.19 (0.38-3.76) |  |  |
|  | Dominant | G/G | 227 (65.80%) | 196 (56.20%) | 1.00 | 0.023 |  |
|  |  | G/A-A/A | 118 (34.20%) | 153 (43.80%) | 1.43 (1.05-1.96) |  |  |
|  | Recessive | G/G-G/A | 339 (98.30%) | 343 (98.30%) | 1.00 | 0.950 |  |
|  |  | A/A | 6 (1.70%) | 6 (1.70%) | 1.04 (0.33-3.27) |  |  |
|  | Overdominant | G/G-A/A | 233 (67.50%) | 202 (57.90%) | 1.00 | 0.022 |  |
|  |  | G/A | 112 (32.50%) | 147 (42.10%) | 1.44 (1.05-1.97) |  |  |
|  | Log-additive | - | - | - | 1.37 (1.02-1.82) | 0.034 |  |

Genotype frequencies in VSD/control were compared using chi-square test;
\*: P value for Hardy-Weinberg equilibrium test in the control subjects.

**Table 3.** Haplotype analysis of the SNPs in the *HAND2* promoter region and the association with VSD risk (Adjusted by age and gender).

| No | rs2276940 | rs2276941 | Freq<br>(Control) | Freq<br>(Case) | Freq<br>(Total) | OR<br>(95 %CI) | P<br>value |
| --- | --- | --- | --- | --- | --- | --- | --- |
| 1 | G | G | 0.8150 | 0.7688 | 0.7918 | 1.00 | - |
| 2 | G | A | 0.1444 | 0.1553 | 0.1499 | 1.16<br>(0.84 - 1.60) | 0.360 |
| 3 | T | A | 0.0353 | 0.0725 | 0.0540 | 2.14 | 0.003 |
|  |  |  |  |  |  | (1.29 - 3.57) |  |
| 4 | T | G | 0.0053 | 0.0035 | 0.0043 | 0.65 | 0.640 |
|  |  |  |  |  |  | (0.11 - 3.96) |  |

### Two SNPs decreased the transcriptional activity of the *HADN2* gene promoter

The influence of the genetic variants on the transcriptional activity of the *HAND2* gene promoter was analyzed using the luciferase reporter gene assay. Expression constructs containing wild-type and variant *HAND2* gene promoters pGL3-WT (wild type), pGL3-173531403T, pGL3-173531377G, pGL3-173531217G, pGL3-173531213G, pGL3-173531210C, pGL3-173531173G, pGL3-173530852G, pGL3-173530848T, pGL3-173530678A, pGL3-173530652A, pGL3-173530503A and pGL3-173530392T, were transfected into HEK-293 and primary NRCMs cells. It should be noted that since the two genetic variants, 173530848 G>T and 173530852 A>G, were presented on the same DNA, we conducted a site-directed mutagenesis for pGL3-173530852G using Quick Mutation™ Site-Directed Mutagenesis Kit (Beyotime). The relative activity of wild-type and variant *HADN2* gene promoters was examined by measuring the dual-luciferase activities. Both in HEK-293 and primary NRCMs cells, the SNP g.173530848 G>T (rs2276940) and g.173531213 C>G (rs138531627) decreased the transcriptional activity of the *HADN2* gene promoter (Fig 3).

**Fig 3.**
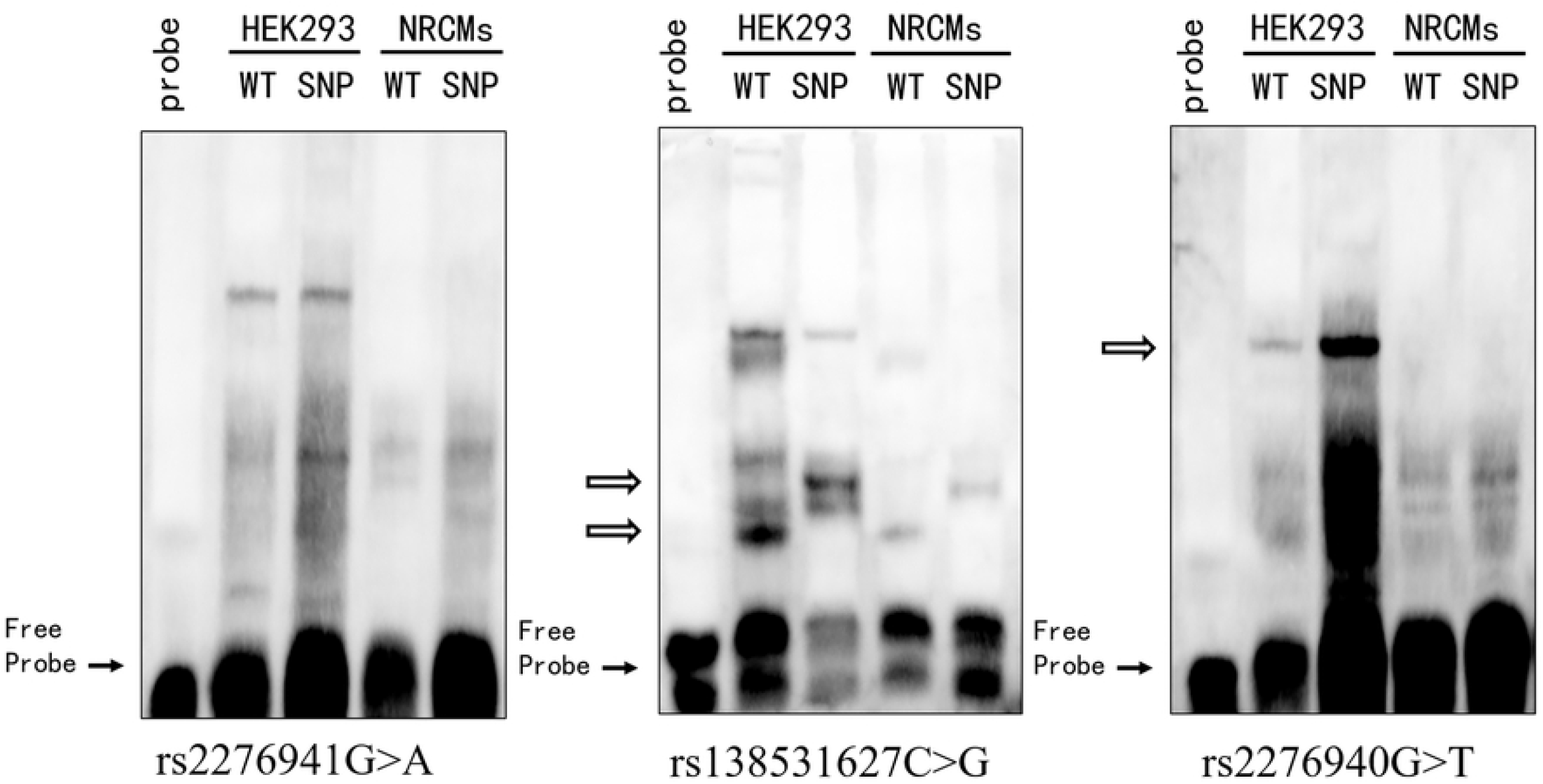
Relative transcriptional activity of wild-type and variant HAND2 gene promoters in HEK-293 and primary NRCMs cells. The data shown are expressed as mean+ SEM. Empty vector pGL3-basic was used as a negative control. The transcriptional activity of the wild-type HAND2 gene promoter was designed as 100%. Lanes 1, pGL3-basic; 2, pGL3-WT; 3, pGL3-173531403T; 4, pGL3-173531377G; 5, pGL3-173531217G; 6, pGL3-173531213G; 7, pGL3-173531210C; 8, pGL3-173531173G; 9, pGL3-173530852G; 10, pGL3-173530848T; 11, pGL3-173530678A; 12, pGL3-173530652A;13, pGL3-173530503A, and 14, pGL3-173530392T. **p< 0.001.

### Prediction of transcription factors

TFs database JASPAR and TRANSFAC were used to predict TFs. The SNP g.173531213 C>G (rs138531627) may abolish the binding sites for *Sp1 transcription factor (Sp1)* and *MYC associated zinc finger protein (MAZ)*, and create the binding sites for *early B-cell factor (EBF) family of transcription factors, NK2 homeobox 1(NKX2-1)* and *BRCA1 DNA repair associated (BRCA1)*. The SNP g.173530848 G>T (rs2276940) may abolish the binding sites for *spermatogenic leucine zipper 1 (SPZ1)*, and enhance the binding affinity for *upstream transcription factor 1 (USF1)* and *neurofibromin 1 (NF1)*. The SNP g.173530503 G>A (rs2276941) may enhance the binding affinity for *forkhead box M1 (FOXM1)* (Table 4).

**Table 4.** Predicted binding sites for transcription factors affected by the SNPs.

| SNP | Change mode | Transcription factors |
| --- | --- | --- |
| g.173531213C>G (rs138531627) | abolish | Sp1; MYC associated zinc finger protein (MAZ) |
|  | create | early B-cell factor (EBF)<br>family of transcription<br>factors; NK2 homeobox<br>1(NKX2-1); BRCA1<br>DNA repair associated<br>(BRCA1) |
| g.173530848G>T<br>(rs2276940) | abolish | spermatogenic leucine<br>zipper 1 (SPZ1) |
|  | modify | upstream transcription<br>factor 1 (USF1);<br>neurofibromin 1 (NF1) |
| g.173530503 G>A<br>(rs2276941) | modify | forkhead box M1<br>(FOXM1) |

### Two SNPs altered the binding of transcription factors

In order to further explore whether the SNPs interfered with the binding of TFs, EMSA was performed with wild-type and variant oligonucleotides (Table 5). The SNP g.173531213 C>G (rs138531627) created a new binding site for TFs in both HEK293 and NRCMs cells, while the SNP g.173530848 G>T (rs2276940) enhanced the binding affinity for TFs only in HEK293 cells (Fig 4).

**Table 5.**
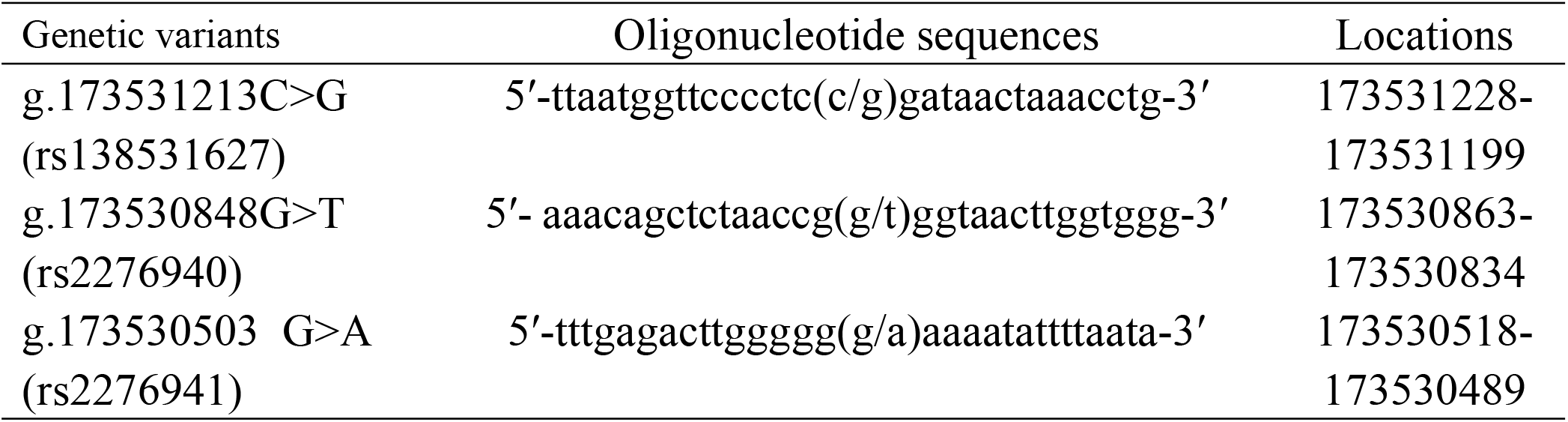
The double-stranded biotinylated oligonucleotides for the EMSA.

**Fig 4.**
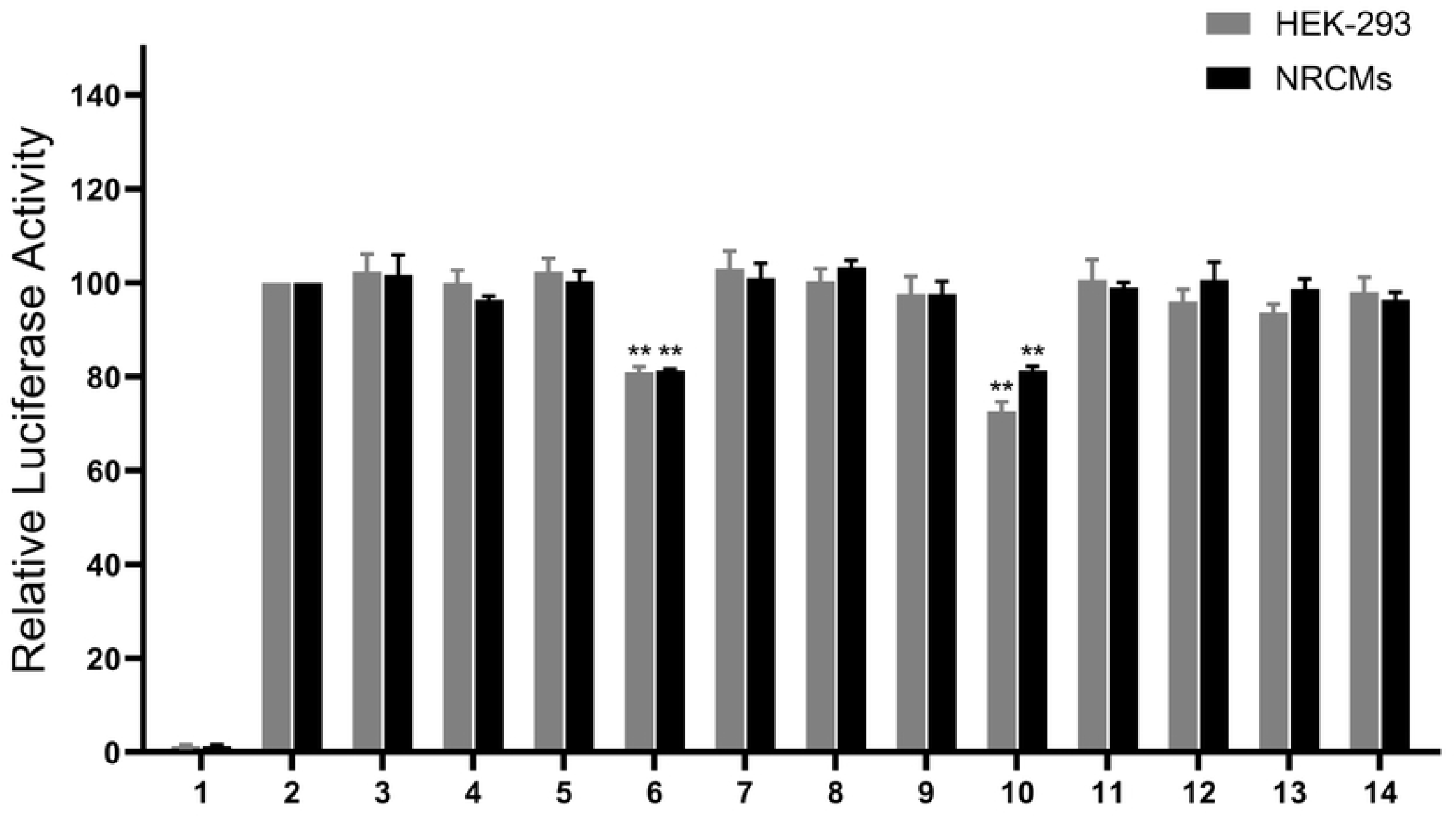
EMSA of biotin-labeled oligonucleotide-containing regulatory variants with nuclear extracts of HEK-293 and NRCMs cells. Free probe was marked at the bottom. The affected binding for an unknown transcription factor was marked with an open arrow. WT, wild type. GV, genetic variants.

## Discussion

The formation of the interventricular septum in humans starts at the end of the fourth week of embryonic development. As the heart develops, the muscular part of the interventricular septum grows upward. By about the eighth week of embryonic development, the interventricular septum is fully formed as the closure of the interventricular foramen[25-28]. Any error during this process may lead to VSD. And *HAND2* is specifically expressed during this process. We therefore hypothesized that the decrease of the *HAND2* promoter transcriptional activity which then induces to the decrease of the *HAND2* expression may lead to VSD.

In the present study, two SNPs [g.173530848 G>T (rs2276940) and g.173530503 G>A (rs2276941)] in the promoter of the *HAND2* gene were revealed to have associations with an increased risk of VSD in the case–control studies. However, the SNP rs2276941 G>A not altered the transcriptional activity of the *HAND2* gene promoter. We speculated that rs2276941 G>A may be in linkage disequilibrium with one SNP which could decrease the transcriptional activity of the *HAND2* gene promoter. Since the promoter fragment we selected was only about 1300bp, this key SNP may have been missed. While the SNP rs2276940 G>T decreased the transcriptional activity of the *HAND2* gene promoter. Therefore, we considered that rs2276940 G>T may play an important role in the development of VSD. Another SNP which was responsible for reducing the transcriptional activity of the *HAND2* gene promoter was g.173531213 C>G (rs138531627). And this SNP was identified only in patients with VSD. Of course, this may be due to the insufficient sample size. Subsequently, we predict the TFs using TFs database JASPAR and TRANSFAC. The result showed that all three above SNPs may change the binding of the TFs. Then we explored the molecular mechanism by conducting EMSA for these SNPs. The results showed that rs138531627 C>G created a new binding site for TFs in both HEK293 and NRCMs cells, while rs2276940 G>T enhanced the binding affinity for TFs only in HEK293 cells. Therefore, we speculated that the reduced transcriptional activity of *HAND2* promoter caused by rs138531627 C>G and rs2276940 G>T may be due to the changes in TFs binding. However, since there is no commercial EMSA antibody, we did not further verify the binding of TFs and these SNPs.

In summary, we found three SNPs that are helpful in identifying the genetic causes of VSD and conducted functional analysis and molecular mechanism study on them. These conclusions have certain reference significance for exploring the molecular genetic mechanism of VSD and clarifying the regulatory network of *HAND2*. However, we still need to expand the sample size and in-depth mechanism research to further verify this conclusion and clarify the relevant molecular mechanism.

## Data Availability

All relevant data are within the manuscript and its Supporting Information files.

